# Psoriatic disease and body composition: a systematic review of the literature

**DOI:** 10.1101/2020.05.18.20104802

**Authors:** T. Blake, N.J. Gullick, C.E. Hutchinson, T.M. Barber

## Abstract

**Background:** Obesity is a leading comorbidity in psoriatic disease, including both psoriasis (Pso) and psoriatic arthritis (PsA), and is associated with adverse metabolic and cardiovascular (CV) outcomes. Anthropometric parameters, such as weight, body mass index (BMI) and waist-to-hip ratio, have been extensively reported in psoriatic disease. However, the associations of body composition and fat distribution with psoriasis have not been fully defined.

**Objectives:** To identify whether patients with psoriatic disease, including psoriatic arthritis, have altered body composition compared with the general population, and to review existing modalities for the assessment of body composition.

**Methods:** Electronic searches of the literature were conducted in PubMed, Medline (Ovid^®^), Embase (Ovid^®^), Cochrane Central Register and Google Scholar. Titles and abstracts were reviewed by two authors independently against a set of prespecified inclusion/exclusion criteria.

**Results:** Twenty-five full text articles met the inclusion criteria and were included in the final narrative analysis. The studies were of heterogeneous design and used a range of objective measures to assess body composition, including simple anthropometric measures, bioimpedance analysis (BIA), dual energy X-ray absorptiometry (DXA) and computed tomography (CT). Few studies met all the quality assessment criteria.

**Conclusions:** Patients with psoriatic disease reveal defined body composition changes that are independent of obesity and the customary metabolic syndrome, including higher overall body fat, visceral fat and sarcopenia. These findings emphasize that patients with psoriatic disease should be screened for abnormal adipose effects beyond their weight and body mass index (BMI). Our findings show that the last decade has seen an exciting expansion of research interest in the development and validation of new modalities for the assessment of body composition. There is no consensus on the optimal assessment method of body composition for this diverse group, hence there is a need for validation of existing modalities and standardization of assessment tools.

## Introduction

Psoriasis (Pso) is an immune-mediated chronic inflammatory disease affecting the skin, entheses and joints, characterized at the skin level by infiltration of immune cells in the dermis and epidermis, vascular proliferation and atypical keratinocyte differentiation. Pathogenesis is complex and thought to result from the interaction between genetic, environmental and immunologic factors; key players in this process are T cells, antigen presenting cells, keratinocytes, Langerhans’ cells, macrophages, natural killer cells, as well as multiple cytokines and growth factors including vascular endothelial growth factor and keratinocyte growth factor [1]. In recent years, the mindset has shifted from one of a Th1-driven immune response with IFN-γ and IL-12 as the signature cytokines to one in which the IL-23/Th17 axis with IL-17, IL-21 and IL-22 plays a more central role [2].

There is increasing recognition that psoriasis is more than skin deep and has important consequences beyond the skin. Proinflammatory molecules released during chronic inflammation are implicated in certain co-morbidities, such as obesity, hypertension, diabetes mellitus, cardiovascular disease and depression. Conversely, epidemiological evidence infers that obesity, via pro-inflammatory pathways, predisposes to both development and progression of psoriasis [3]. This association is shared with metabolic syndrome (MetS), not least the increased prevalence of cardiovascular risk factors and the ensuing cardiovascular morbidity [4–8]. Recent studies have suggested that adipokines, such as leptin, adiponectin and resistin, produced by adipocytes and dysregulated in obesity and MetS, are the linchpins of this metabolic association in psoriatic disease. They have been shown to contribute independently to the adverse cardiovascular outcomes in patients with Pso and can be viewed as biomarkers of obesity-related inflammation and cardiovascular risk (9). With this is mind, one should consider adipose tissue as an endocrine organ that has the capabilities, through local and systemic factors, to induce a low-level inflammatory state. Moreover, specific IL-17-secreting Th17 cells and IL-22-secreting Th22 cells have been seen to infiltrate the adipose tissue and represent local mediators of inflammation and insulin resistance, something that is being studied in more detail.

Sustained inflammation due to psoriatic disease also leads to loss of muscle mass and muscle weakness termed sarcopenia; however, the reasons for these muscle changes in the context of inflammation are multifactorial and difficult to define. External factors, such as aging, decreased physical activity secondary to stiffness and pain, hormonal changes and disturbances in protein metabolism are likely to exert considerable influence on this process.

Despite this knowledge about the associations between psoriatic disease, adipogenesis and metabolic dysfunction, there is less emphasis on body mass alterations and distribution between separate internal compartments: fat-free tissue (lean body mass), extracellular water and adipose tissue. Body composition, a measure of lean and fat mass proportions, provides a useful indicator of metabolic health [10], and its measurement in psoriatic disease can provide a useful insight into cardiometabolic risk, something that could well influence patient management.

Our principal aim was to determine evidence for association between psoriasis and abnormal patterns of fat (including preponderance of visceral fat content) and muscle distribution. We also explored and compared different modalities used for assessment of fat distribution. We hope that this analysis may guide future clinical decision making with respect to risk assessment, screening and management of psoriatic patients in day-to-day practice, encouraging more individualized care and leading to better patient outcomes.

## Methods

This systematic review was performed following methodology recommended by the Cochrane Collaboration and is reported according to the Preferred Reporting Items for Systematic Reviews and Meta-analyses (PRISMA) guidelines [11, 12].

### Eligibility criteria

Studies were included where the primary aim was to elucidate an association between psoriasis or psoriatic arthritis and abnormal body composition. Studies were not required to include a comparator group. The following inclusion and exclusion criteria were applied.

### Inclusion criteria

Types of studies

- Publication date 1999 (inclusive) - present
- Studies from any geographical location
- English language
- Published studies (including conference papers)
- Grey literature (not published in a peer-reviewed journal) including dissertations/theses
- Any quantitative study (RCT, non-RCT, observational, cohort, case control)
- Studies using qualitative methods of analysis (to describe patterns or themes raised by studies) seeking to understand body composition phenotypes of psoriatic disease. This includes original qualitative studies, studies involving secondary analysis of data, and a qualitative study as part of a mixed methods study e.g. the study also has a quantitative component
- The reference lists of the final articles included for full data extraction were hand searched for further relevant articles.

Types of participants

- Adults (>18 years)
- Diagnosed with psoriasis, psoriatic disease or psoriatic arthropathy/arthritis
- Being treated in any ‘usual care’ setting: primary; secondary; tertiary care, e.g. in the hospital, hospice, community, home or rehabilitation
- Receiving care typical for that geographical location

Types of outcome measures

- Understanding and learning about body composition and metabolic phenotypes including measures of body fat and lean mass in men and women with psoriatic disease
- To explore the metabolic, anthropometric and biochemical indices of psoriatic disease

Exclusion criteria:

- Non-English language
- Published pre-1999
- Any study where quantitative or qualitative data are not analyzed i.e. uninterpreted data; case reports; any editorial, meta-analysis or review (systematic, narrative, qualitative)
- Treatment guidelines and pathways
- Commentary articles, written to convey opinion or stimulate research /discussion, with no research component

Types of participants

- Children or Young adults (<18 years)
- Diagnosis of Spondyloarthritis or Spondyloarthropathy, Ankylosing Spondylitis, other immune-mediated inflammatory diseases; studies that do not focus on psoriatic disease per se

Types of outcome measures

- Anything that is not concerned with demonstrating either a metabolic, structural or functional relationship to psoriatic disease; studies focusing on individual components of metabolic syndrome (diabetes mellitus, hypertension, hyperlipidemia or fatty liver disease) and any association with psoriatic disease

### Databases and searches

The search strategy was developed by two of the authors (TB and NG) and a librarian. Database searches were performed in PubMed, Medline (Ovid^®^), Embase (Ovid^®^), Cochrane Central Register and Google Scholar for reports published between 1999 to November 2019 using a sensitive methodological filter for studies. Search terms and criteria are shown in Table 1. Search results were combined into a single Endnote file and duplicates removed.

**Table 1.** Search terms.

|  |
| --- |
| **Psoriatic disease** |
| psoriatic arthritis.mp. or exp Arthritis, Psoriatic/<br>psoriatic arthropathy.mp. or exp Arthritis, Psoriatic/<br>exp Psoriasis/ or psoriatic disease.mp.<br>exp Spondylarthritis/ or Spondyloarthropathies/ or spondyloarthritis.mp. |
| **Metabolic composition** |
| metabolic syndrome.mp. or exp Metabolic Syndrome/<br>exp Body Weight/ or metabolic syndrome.mp. or exp Blood Glucose/ or exp Obesity/<br>or exp Lipids/ or exp Insulin/ or exp Metabolic Syndrome/ or exp Glucose Tolerance<br>Test/<br>body composition.mp. or exp Body Composition/<br>exp Exercise/ or sarcopenia.mp. or exp Muscle, Skeletal/ or exp SARCOPENIA/ or<br>exp Muscular Atrophy/ or exp Muscle Proteins/<br>sarcopaenia.mp. |
| **Limits** |
| English language<br>Humans<br>Last 15 years of publication |

### Study selection

Two authors (TB, NG) independently reviewed all titles/abstracts in the web-based software platform Covidence [13] and selected articles for full-text review. Discrepancies were resolved by consensus.

### Data extraction and quality assessment

The data extraction tool was designed by TB and included a Quality Assessment incorporating a Critical Appraisal Skills Programme (CASP) checklist [14] specific for the study design. The checklists included criteria for information and selection bias addressing the following domains: participants, controls, measurement of variables, statistical power, confounding factors and applicability of results. All included studies were appraised with the Risk Of Bias In Non-randomized Studies of Interventions assessment tool (ROBINS-I) [15].

## Results

### Search results

The initial search yielded 2202 titles. After removing duplicates, 1848 titles and abstracts were screened, then 238 retained for full text review. A further 213 studies were excluded due to: incorrect outcomes (197), incorrect study design (9), duplicate material (3), incorrect intervention (3) and non-English language (1). A total of 25 studies from 25 publications met the inclusion criteria (Figure 1) with differing methodology: 1 randomized control trial, 1 case-control study, 19 cross-sectional and 4 prospective cohorts. A search of the grey literature (the first 100 articles sorted by relevance on Google Scholar) retrieved no additional studies. A summary of the included studies is shown in Table 2.

**Figure 1.**
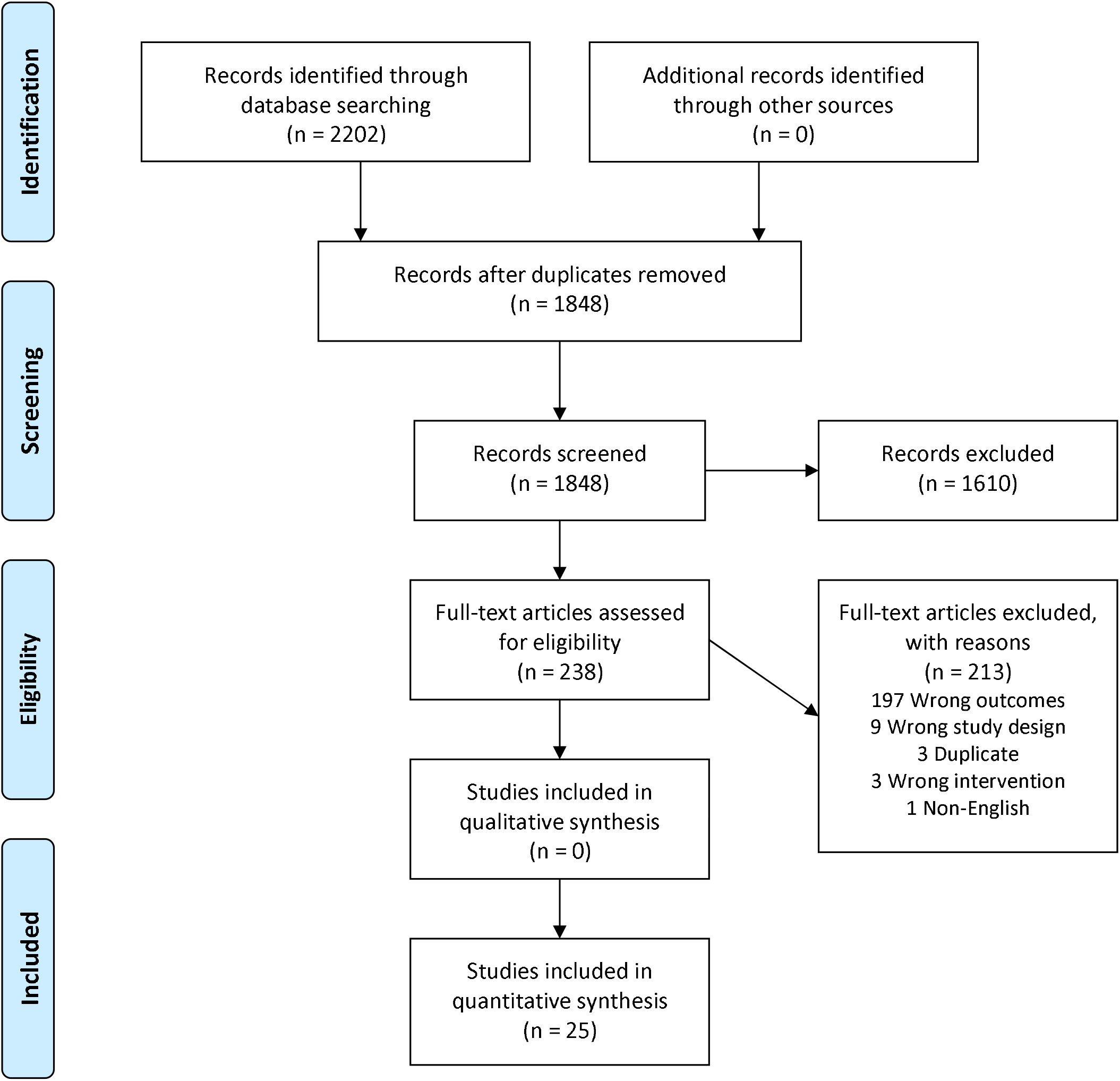
PRISMA 2009 Flow Diagram.

**Table 2.**
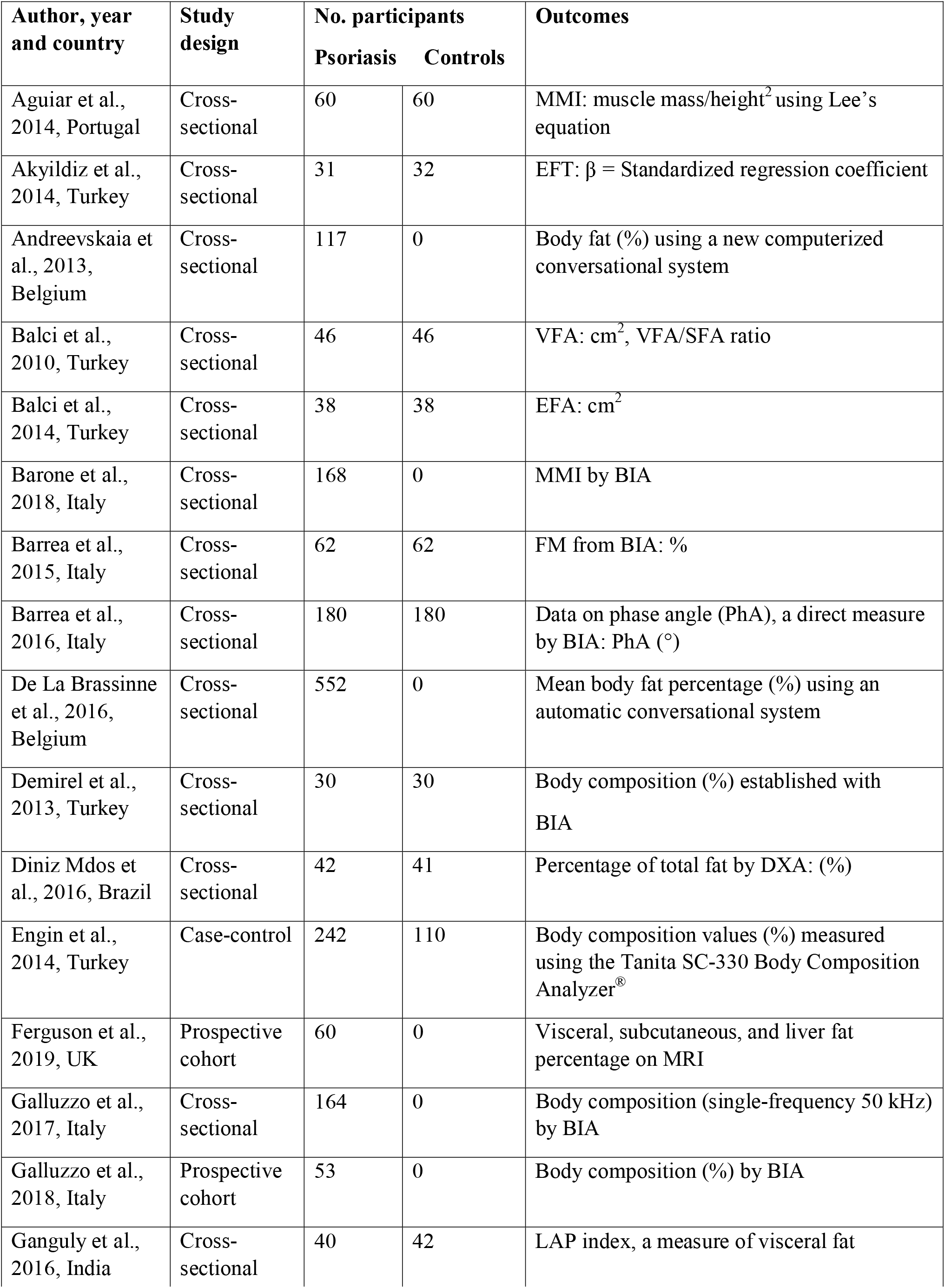
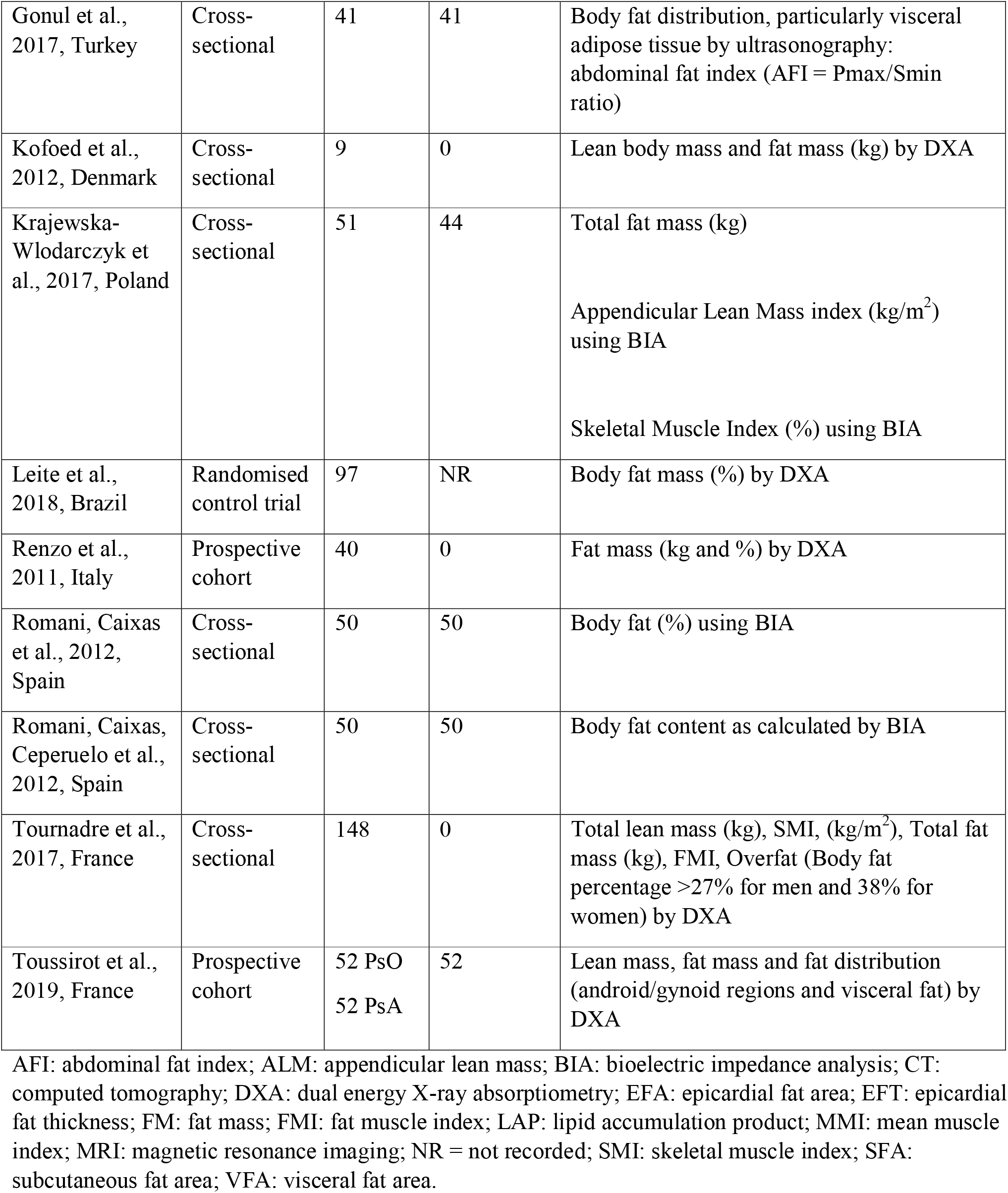
Meta-summary of included studies.

### Population sampled

Twenty-five studies represented a population of 2468 psoriatic patients from 10 countries: Belgium, Brazil, Denmark, France, Italy, Poland, Portugal, Spain, Turkey and UK. Demographics are shown in Table 3. Studies did not differentiate outcomes according to subtypes of psoriatic disease, however five of the studies focused purely on psoriatic arthritis [16–20]. Only three studies [19–21] reported racial and ethnic group of participants. A control group was assessed in 10 studies [16, 18, 19, 21, 23–8], and this information was missing in another study [17]. None of the studies reported socio-economic status including employment or educational attainment of subjects.

**Table 3.** Demographic data of populations.

| **Author, year and country** | **Age of Psoriasis group, years (mean ± SD unless stated)** | **Sex, female %** | **Ethnicity** | **Duration, years (mean ± SD unless stated)** | **Disease type** | **Participant selection** |
| --- | --- | --- | --- | --- | --- | --- |
| Aguiar et al., 2014, Portugal | 45.5 ± 13.4 | 51.7 | Caucasian | 10.9 ± 11.6 | PsA | PsA according to CASPAR criteria. Controls recruited from primary healthcare. |
| Akyildiz et al., 2014, Turkey | 42.0 ± 11.1 | 54.8 | ND | 17.1 ± 10.5 | PsO | Subjects attending the outpatient dermatology clinic. Clinical examination in keeping with psoriasis. |
| Andreevskai a et al., 2013, Belgium | 48.8 ± 14.9 | 49.5 | ND | ND | PsO | Subjects attending a new outpatient psoriasis center recruited at random of their consultation. |
| Balci et al., 2010, Turkey | 39.5 ± 14.2 | 37.0 | ND | 11.0 ± 8.3 | PsO | Patients attending outpatient dermatology clinic. Controls age and sex matched. |
| Balci et al., 2014, Turkey | 42.2 ± 15.0 | 31.6 | ND | 14.9 ± 9.5 | PsO | Patients attending outpatient dermatology clinic. Controls age and sex matched. |
| Barone et al., 2018, Italy | 55.3 ± 9.1 | 84.2 | ND | 11.1 ± 8.1 | PsA | Patients with a clinical diagnosis of RA, SA, and PsA enrolled |
| Barrea et al., 2015, Italy | 50.2 ± 10.5 | 30.0 | ND | ND | PsO | Patients with a diagnosis of mild-to-severe psoriasis lasting for at least 6 months were enrolled, while patients with pustular, erythrodermic or arthropathic psoriasis or receiving any systemic treatment for psoriasis including acitretin, ciclosporin, methotrexate, phototherapy or biologics for at least 3 months were excluded. |
| Barrea et al., 2016, Italy | 50.0<br>(21.0 – 65.0) | 29.0 | Caucasian | ND | PsO | Patients attending a university hospital dermatology clinic. Controls matched based on age, sex and BMI. |
| De La Brassinne et al., 2016, Belgium | 47.68 ± 16.12 | 45.7 | ND | ND | PsO | Subjects attending a new outpatient psoriasis center recruited at random of their consultation. |
| Demirel et al., 2013, Turkey | 37.8 ± 10.6 | ND | ND | ND | PsO | Patients previously diagnosed with psoriasis<br>by a trained dermatologist using biopsy. All patients defined as having mild to moderate psoriasis and receiving only topical therapy. Moderate psoriasis identified by PASI >10. |
| Diniz Mdos et al., 2016, Brazil | 47.0 | 40.5 | ND | 47.0 | PsO | Patients with psoriasis and without joint complaints, adult patients with clinical and/or histopathological diagnosis of psoriasis, without joint complaints, assisted at dermatology outpatient clinic. The control group comprised voluntary patients without psoriasis or any inflammatory disease, matched by sex and age. |
| Engin et al., 2014, Turkey | 42.7 | 50.0 | ND | 13.6 ± 8.8 | PsO | University psoriasis clinic. The diagnosis of psoriasis was based on clinical examination or histopathological examination in all patients. |
| Ferguson et al., 2019, UK | 55.0<br>(43, 62 IQR) | 63.0 | ND | 8.0<br>(2, 12.2 IQR) | PsA | Prospective, open label study of adults receiving apremilast as part of routine care for PsA and/or psoriasis. |
| Galluzzo et al., 2017, Italy | 48.6 ± 16.5 | 36.6 | ND | ND | PsO | Chronic plaque psoriasis patients, naive to biological drugs, treated with ustekinumab in the University dermatology clinic. |
| Galluzzo et al., 2018, Italy | 32.1 ± 13.6 | 24.5 | Caucasian | 17.1 ± 11.5 | PsO | Chronic plaque psoriasis patients, naive to biological drugs, treated with ustekinumab in the |
|  |  |  |  |  |  | University dermatology clinic. |
| Ganguly et al., 2016, India | 44.8 ± 12.8 | ND | ND | 0 - 20 | PsO | Patients with chronic plaque psoriasis attending the dermatology outpatient clinic. Healthy controls who came for a routine health check to our hospital were included in the study. |
| Gonul et al., 2017, Turkey | 43.2 ± 15.6 | 46.3 | ND | 14.6 ± 12.1 | PsO | Patients with clinically and histopathologically diagnosed psoriasis, patients had been receiving systemic or topical treatment for the last month. Controls recruited from among patients with warts, tinea pedis, or melanocytic nevi without any inflammatory disease, and matched for age, sex and BMI. |
| Kofoed et al., 2012, Denmark | 33.0 ± 8.6 | ND | ND | 15.9 ± 4.4 | PsO | Eligible participants were anti-TNF-naïve patients with psoriasis recalcitrant to other systemic treatments and UV-B therapy. They also had a PASI or DLQI of >10. |
| Krajewska-Włodarczyk et al., 2017, Poland | 65.6 ± 5.9 | 100 | ND | PsA 11.1 ± 8.9<br>PsO 22.9 ± 12.9 | PsA/PsO | Women diagnosed with PsA, treated at the rheumatology or dermatology clinic of the municipal hospital. The diagnosis of PsA was determined based on the CASPAR criteria. At least 12 months had passed since the last menstrual period. |
| Leite et al., 2018, Brazil | ND | ND | ND | ND | PsA | ND |
| Renzo et al., 2011, Italy | PsA 42.2 ± 8.9<br>PsO 36.8 ± 8.4 | ND | ND | PsA 14.1 ± 11.2<br>PsO 15.0 ± 9.3 | PsA/PsO | Patients divided into two groups: 20 patients with PsO and 20 with PsA. Twelve patients received infliximab and 28 received etanercept. |
| Romani, Caixas et al., 2012, Spain | 46.4 ± 17.3 | 61.3 | ND | ND | PsO | PASI ≥ 10 evaluated by the same dermatologist. No systemic anti-psoriatic therapy (acitretin, ciclosporin, methotrexate or biologics), natural or artificial UV sources during the month prior to inclusion. |
| Romani, Caixas, Ceperuelo et al., 2012, Spain | ND | ND | ND | ND | PsO | Patients eligible for narrow-band UVB phototherapy recruited excluding the months of high solar irradiance. Controls without psoriasis matched to the patients' characteristics. Patients with PsA were excluded, along with patients and controls with a list of chronic inflammatory diseases. |
| Tournadre et al., 2017, France | 54.6 ± 11.0 | 63 | ND | 5.5 ± 6.8 | PsA | ND |
| Toussirot et al., 2019, France | PsA 52.5 ± 11.7<br>PsO 50.5 ± 12.8 | 51.2<br>26.9 | ND | PsA 9.1 ± 6.7<br>PsA 18.1 ± 13.8 | PsA/PsO | Patients with Pso (plaque psoriasis) or PsA (CASPAR criteria) were evaluated. Each patient was paired to a control subject, recruited in the same outpatient population, and matched for sex, age and BMI. |

ND: not defined.

### Clinical anthropometric assessment

All studies included clinical anthropometric data to varying degrees for overall measurement of participant shape and size. In all but one study [17], participants’ weight, height and BMI were measured. Several studies measured waist-to-hip ratio [27, 29–31], and one study supplemented this with skinfold thickness and abdominal circumference [29]. Only two studies commented on the attire of recruited subjects and attempted to reduce confounding through clothing by stipulating a dress code of light clothes and no shoes [22, 32]. These groups were assessed by a standard operator in a consistent manner. Very few studies incorporated participant information on smoking, alcohol, nutrition and physical activity, all of which are known to influence human metabolism [19, 22, 27, 32].

### Body composition

In general, 24 studies confirmed discrete biological and body composition changes in patients with psoriatic disease, which correlated positively with other indicators of metabolic syndrome, including waist circumference, waist-to-hip ratio, weight, BMI, plasma concentrations of low-density lipoprotein (LDL)-cholesterol, leptin and apolipoprotein-B (apo-B).

One study failed to reveal statistically significant differences between psoriatic patients and controls with respect to maximal aerobic capacity, resting metabolic rate, pulmonary function tests, body fatness, body fat distributions and quality of life [29]. Moreover, not all studies found that these changes were correlated to either skin or joint disease activity [17, 33–35].

Three studies focused on muscle mass and reported levels of sarcopenia [16, 19, 20]. Aguiar et al evaluated muscle mass by way of a muscle mass index (muscle/height^2^) and demonstrated reductions for spondyloarthritis with no significant variation between psoriatic arthritis and ankylosing spondylitis patients; however, their findings were not correlated with disease duration activity, function or radiological indices. A more recent study assessed sarcopenia and presarcopenia on the basis of European Working Group On Sarcopenia in Older People (EWGSOP) criteria and by using defined MMI cut-offs, and identified sarcopenia in 20.0% and presarcopenia in 25.7% of PsA patients [19].

### Measurement techniques

Ten techniques were used to describe body composition: BIA 10, DXA 6, CT 2, ultrasonography 1, transthoracic echocardiography 1 and other techniques 5 (including novel automated systems for measurement). Body composition outcomes by modality are shown in table 4. Four studies [21, 22, 25, 32] measured Phase Angle (PhA), a direct measure of BIA, which researchers found was lower in the psoriatic patients. Barrea et al [22]. discovered that PhA was inversely associated with disease severity measured by Psoriasis Area Severity Index (PASI) and Dermatology Life Quality Index (DLQI); this was also found to be independent of BMI (*P* < 0.001), although this study included low numbers of subjects.

**Table 4.**
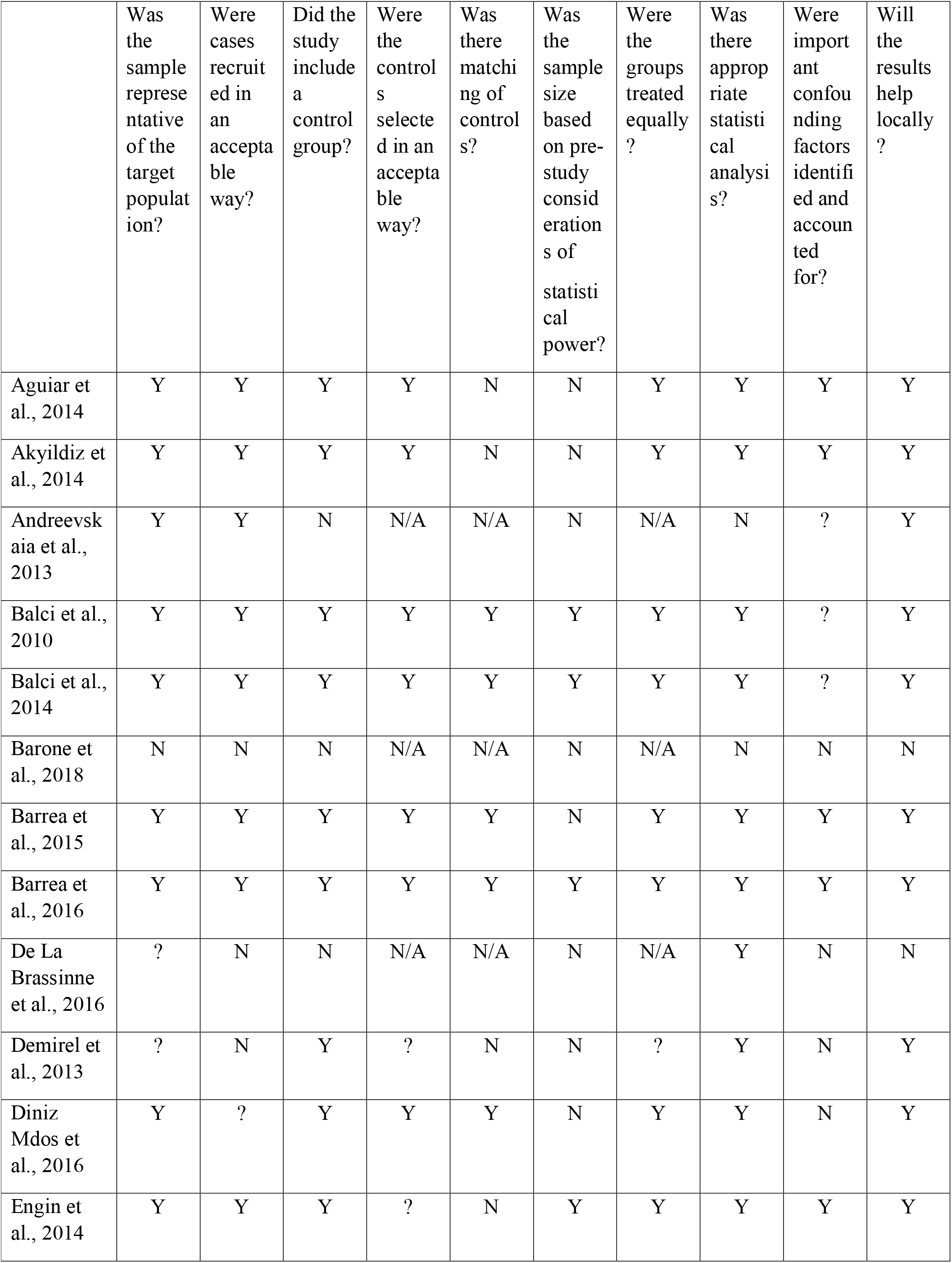
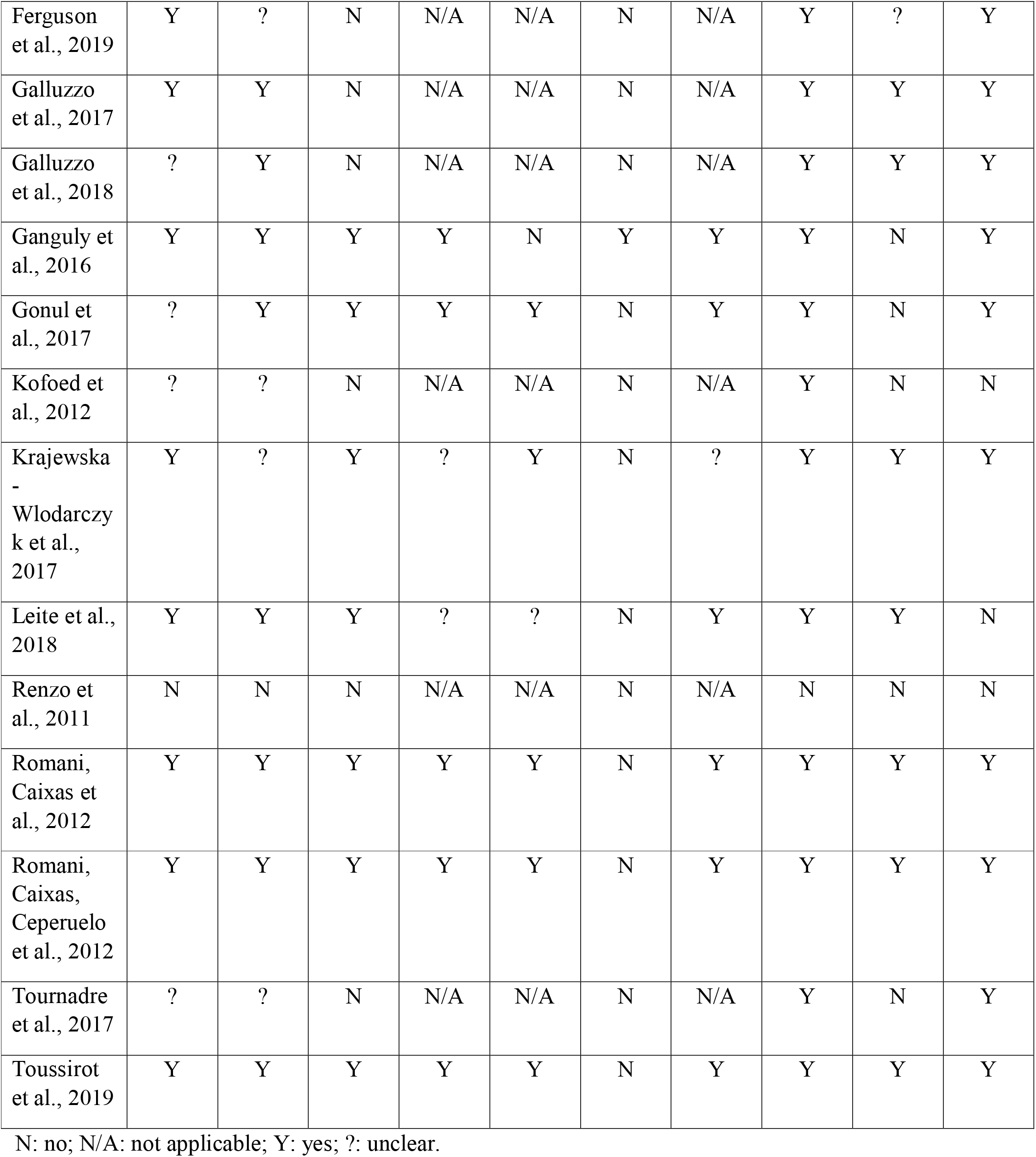
Risk of bias assessment.

**Table 5.**
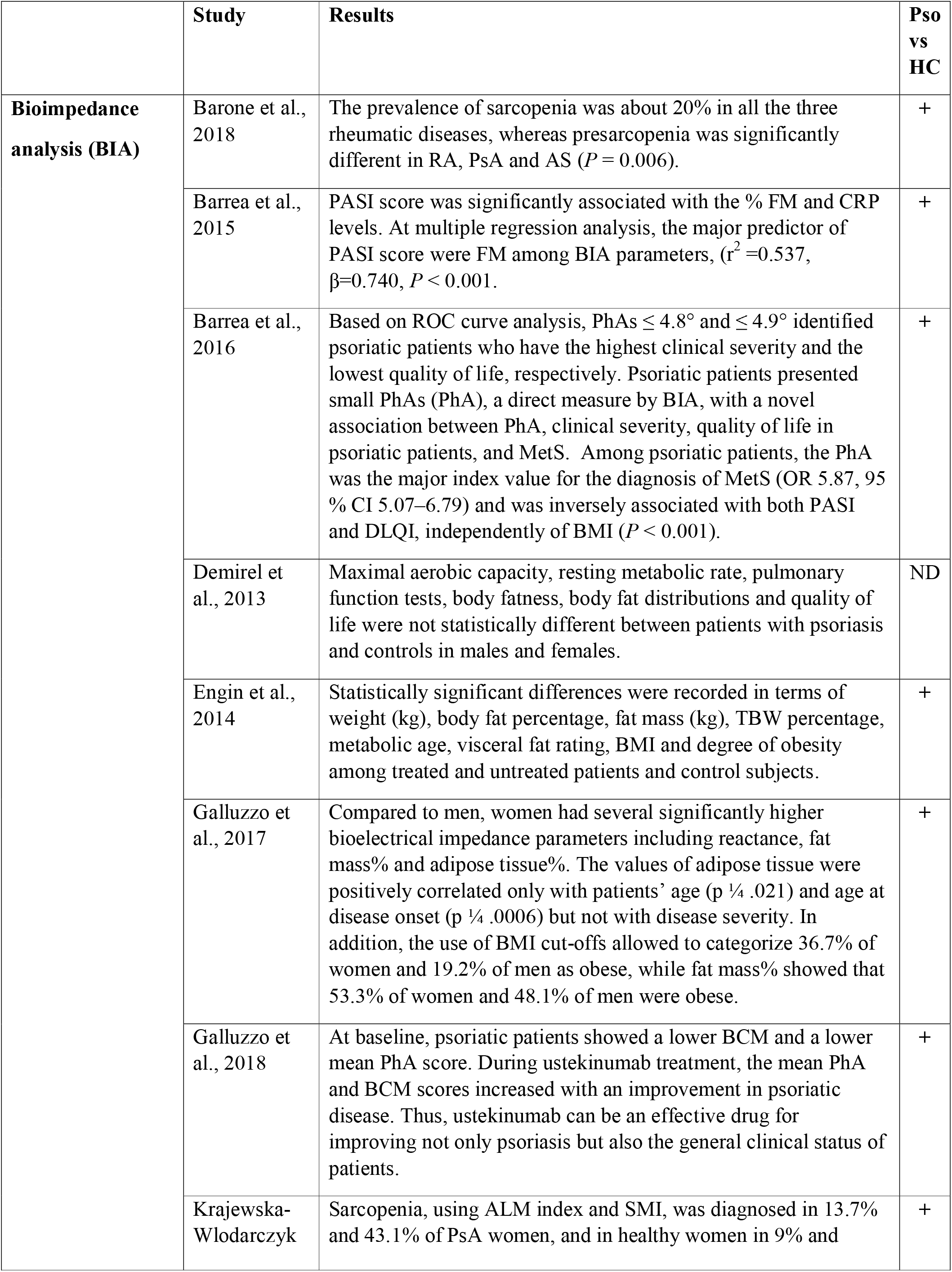
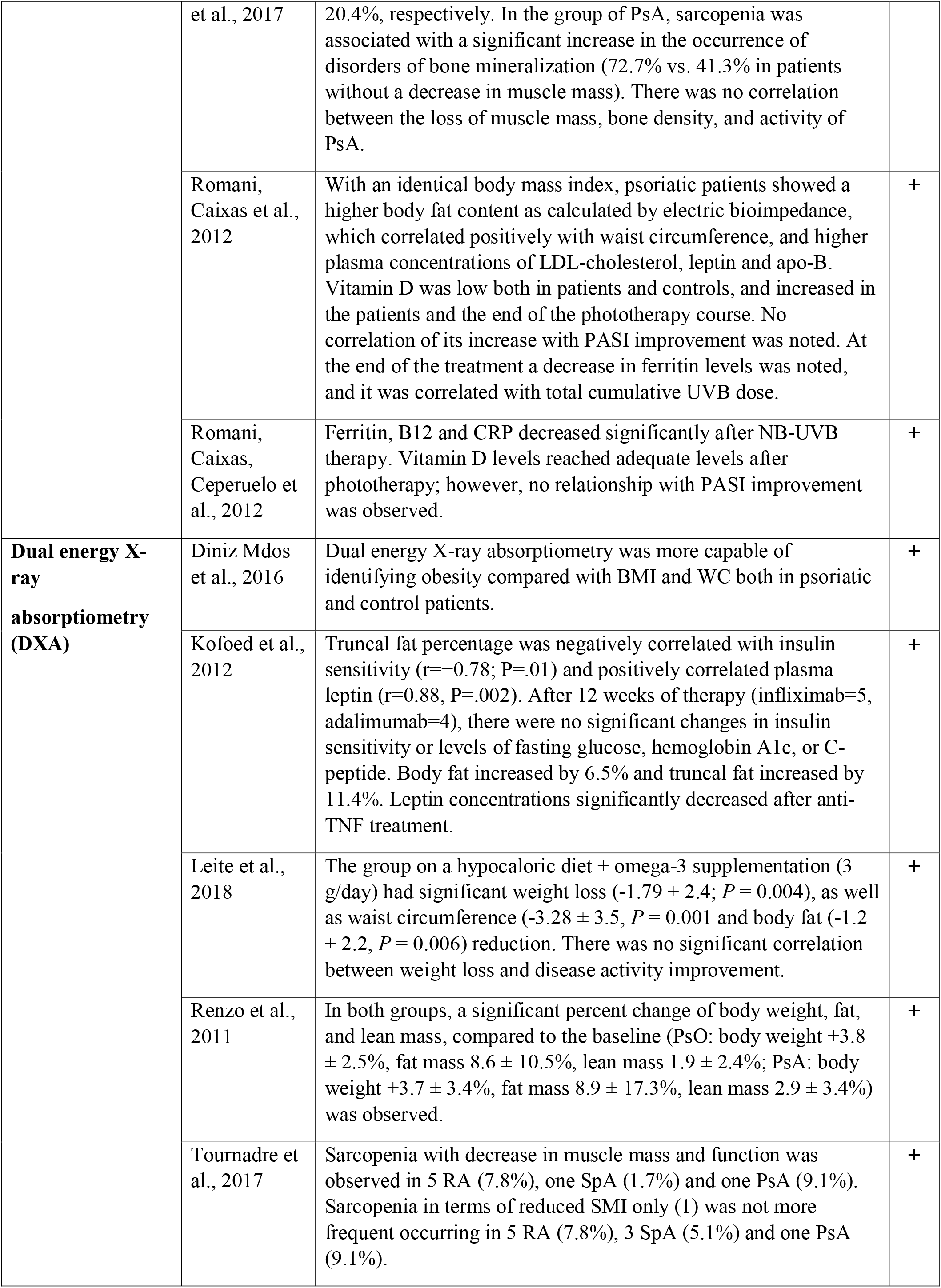
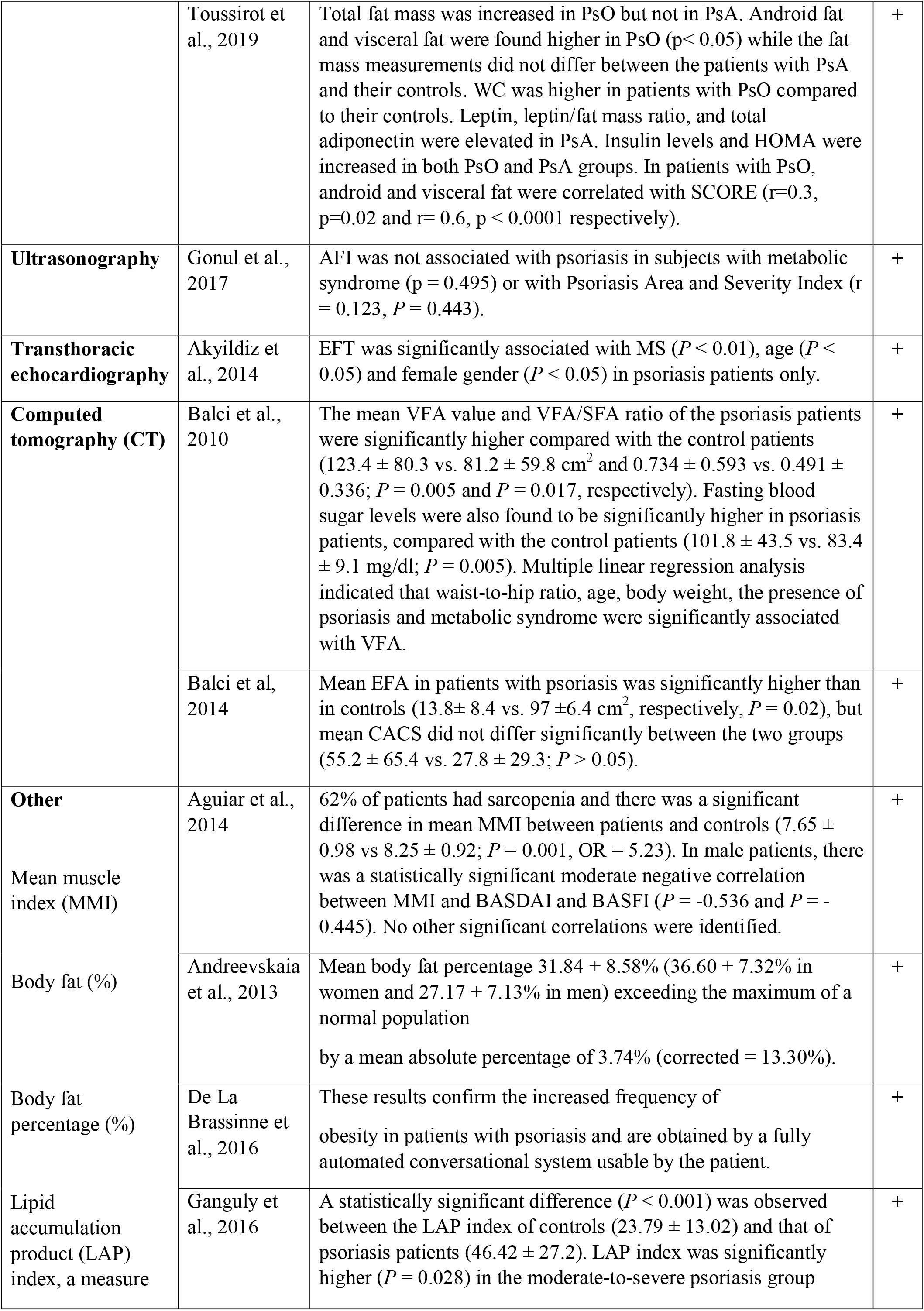
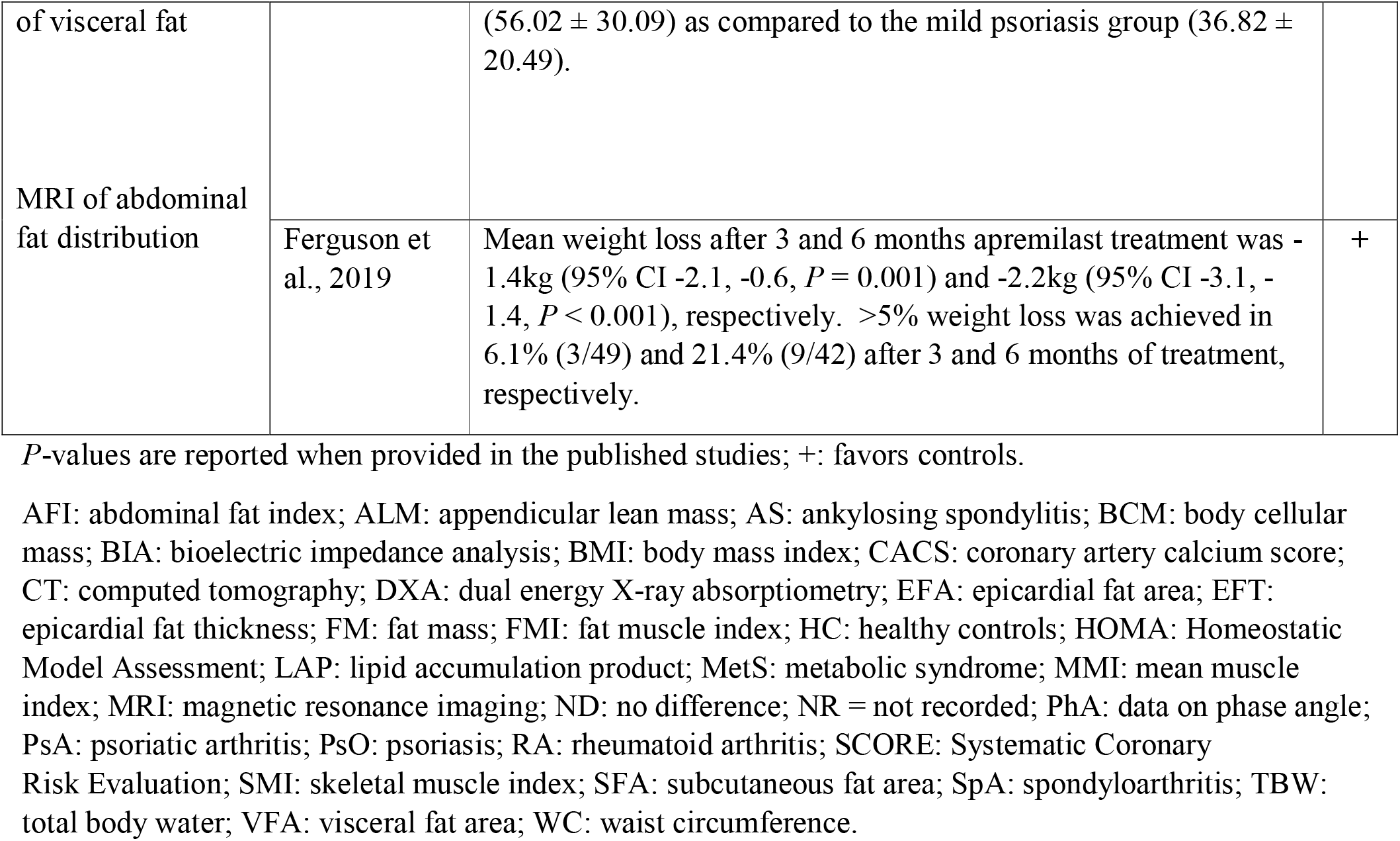
Body composition outcomes by modality.

The following devices were used to derive body composition data for analysis in the 10 included BIA studies: BIA 101, Akern srl, Pontassieve, Florence, Italy [19]; single-frequency 50 kHz BIA 101 RJL, Akern Bioresearch, Florence [21, 22, 25, 32]; Bodystat 1500, Bodystat Ltd., Douglas, Isle of Man, UK [28], Tanita SC-330 Body Composition Analyzer, Tanita Corp., Tokyo, Japan [36], InBody 170, Biospace, South Korea [33]; Tanita TBF300, Tanita Corporation, Tokyo, Japan [34, 35].

All studies bar one [29] demonstrated less favorable body composition data in psoriasis patients compared to controls.

### Body composition associations

Balci et al. [30] identified increased visceral fat area (VFA) using CT in psoriasis patients vs. controls. Multiple linear regression analysis in all study subjects indicated that VFA was significantly associated with waist:hip ratio, age, body weight and presence of metabolic syndrome, though not PASI score, duration or type of disease, smoking habit and therapies. A second study from this group studied epicardial fat [31], in which epicardial fat area (EFA) and coronary artery calcium scoring (CACS) were measured in patients with psoriasis and controls: EFA was significantly associated with CACS, waist circumference and age in the psoriasis patients only. No clinical features or laboratory findings were coupled to EFA. Barone et al. [19], showed age, CRP and disability were associated with sarcopenia, whereas the type of rheumatic disease (RA, PsA or AS), gender, calorie and protein intake, physical activity level, biologic treatment, duration of disease and ESR were not associated with an increased risk of sarcopenia.

### Effect of treatments

Six studies reported on cardiometabolic and body composition improvements on patient profiles after receiving specific interventions: hypocaloric diet with omega-3 supplementation [17], phosphodiesterase-4 (PDE4) inhibition with apremilast [18], narrowband ultraviolet (NB-UVB) therapy [34], anti-TNFα administration with infliximab or etanercept [26], anti-TNFα administration with infliximab or adalimumab [26] and anti-IL-12/23 administration with ustekinumab [21].

### Assessment of bias

The risk of bias quality assessment of included studies is presented in Table 3. Six of the studies, whilst identifying as case-control, were reassigned as cross-sectional design on account of their methodology [20, 22, 35, 37–39]. Nine studies did not include a control group [16, 18, 19, 21, 23–27]. Several studies did not fully specify how their participants were recruited [19, 24, 27, 29] and there was uncertainty about this information from an additional four studies [16, 18, 26, 33]. The sample size was small (< 40) for a number of studies [26, 29, 31, 37]. Overall, four studies were regarded to be at high risk of bias [15, 18, 23, 26], two at medium risk of bias [26, 29] and 18 at low risk of bias [18, 20–23, 25, 28, 30, 31, 33–41] using the ROBINS-I tool.

## Discussion

To our knowledge, this is the first systematic review examining the relationship between psoriatic disease and whole-body composition as a distinct entity from metabolic syndrome. Strengths of the review include use of a high degree of rigor in its search strategy and screening procedures, using relevant standards for performing systematic reviews. We tried to reflect the complete picture of both published and unpublished literature on this topic. The last decade has seen an exciting expansion of interest in the development and validation of new modalities for the assessment of body composition. Our study provides evidence for a relationship between certain body composition phenotypes and the occurrence of psoriasis, including higher overall body fat, visceral fat and sarcopenia that is similar, yet distinct, from the metabolic syndrome.

Several aspects of body composition, specifically the amount and distribution of body fat and lean mass, are now understood to be important health outcomes in adults and should form an important part of the ongoing clinical assessment of patients with psoriasis. However, the issue of whether body compartment distribution is a result of severe psoriasis or a causative factor in its development remains contentious. It is likely that novel systems will eventually supplement less sophisticated bedside measurements and influence key aspects of risk assessment, prognostication and management.

We found an increased prevalence of body composition derangements in psoriatic patients compared with controls. As expected, parameters associated with obesity, such as weight, body fat percentage, fat mass, and degree of obesity, were higher in the psoriasis groups than in the control groups, irrespective of therapy.

Such an association between obesity and psoriasis has been well documented, first described in 1986 [42]. Traditional epidemiological studies have focused on weight or BMI to define obesity rather than altered body composition. We found conflicting data on the association between psoriasis severity, such as PASI, and body composition parameters, indicating that a causal link is not definitive. Previous studies have alluded to a dose-response relationship between psoriasis severity and metabolic syndrome [43], supported by translational studies showing T-helper cell cytokine upregulation in the blood and skin of psoriasis patients, leading to effects on lipid metabolism and insulin resistance [44].

In this review, treatment with anti-IL-12/23 or PDE4 inhibitors was associated with more favorable body composition profiles than anti-TNFα treatments, findings which mirror previous observations of increases in BMI seen with this drug class [45, 46]. IL-17, one of the key proinflammatory cytokines in psoriasis, mechanistically links inflammation with insulin resistance and adipocyte dysfunction [47]. IL-17A producing cells are thought to be pathogenic in driving inflammation in obesity and progression of obesity-related inflammatory diseases, suggesting that causality between psoriasis and adipogenesis is likely to be bidirectional [48]. From this perspective, there are likely to be therapeutic implications of targeting proinflammatory factors like IL-17 or IL-12/23 in metabolic dysfunction associated with psoriatic disease.

Due to its non-invasiveness, low cost and portability, it is easy to appreciate how BIA was adopted by most researchers. The technique relies on the assumption that the volume of fat-free tissue will be proportional to the electrical conductivity of the body. It employs a small electric current to measure the resistance and reactance at difference frequencies against various tissues in the body e.g. lipid has a high resistance to the flow of current, therefore shows a high impedance reading, whereas muscle, which stores most of our body water, has lower impedance. BIA assessment tools have been considered a promising approach for the quantitative measurement of tissue characteristics over time, as well as demonstrating the direct relativity between fluctuations in body composition and prognosis, clinical condition and quality of life [49]. The technique offers reliable data on body composition provided that suitable (i.e. age-, sex- and population-specific) equations for the calculation of body compartments are applied [50]. However, a major limitation of this technique pertains to measurement discrepancies between devices from different manufacturers and the lack of internationally recognized standard reference values.

PhA is thought to be one of the most clinically relevant parameters of BIA. It is defined as the ratio of resistance (intracellular and extracellular resistance) to reactance (cell membrane-specific resistance), expressed as an angle. It is considered an indicator of cellular health, where higher values reflect cell membrane integrity and better cell function. In healthy populations, increasing age bestows a lower PhA due to a reduction in reactance and a parallel loss of muscle mass and an increase in resistance due to the declining proportion of body water at the expense of fat mass. In disease, PhA is often reduced because of infection, inflammation or disease-specific determinants [51]. Recent studies have reported that PhA in humans follows a linear relationship with cellular health and can be considered a prognostic tool in certain medical disorders, including cancer, cirrhosis and diabetes mellitus [22, 52–56]. It is important to note that not all BIA devices can detect phase-sensitive impedance variation that can be used for assessment of phase angle.

It is important to recognize that there is no single measurement method of body composition that allows for the delineation of all tissues and organs and there are pros and cons of all techniques. The seemingly unsophisticated measurements of skin thickness, BMI and waist circumference can provide simple longitudinal assessments of fatness and metabolic risk despite their poor accuracy and inability to differentiate fat and lean masses. The value of any approach in supporting clinical practice is enhanced by the availability of reference data. Recent developments include MRI for fat distribution.

### Limitations

This systematic review should be interpreted in the context of the reported studies which were heterogeneous in several aspects. Firstly, the observational studies recruited different extents and subtypes of psoriatic disease, some with associated arthritis, and measured different aspects of body composition, making definitive conclusions problematic. Secondly, there was poor matching of patients and controls across all studies and little consideration for the potential confounding effects of key determinants of metabolism, e.g. physical activity, age and smoking. Finally, BIA-estimated percentage of body fat varies greatly with population and age and is directly and closely related to various health outcomes such as cardiovascular diseases. Despite its prognostic potential, BIA has not been validated in population studies or clinical practice due to lack of normal population reference limits for comparison and is also influenced by other factors such as age, sex and race [57].

## Data Availability

All data generated or analyzed during this study are included in this published article.

## Further considerations

We suggest that body composition indices should be analyzed in more detail with a broader range of techniques and imaging systems across the clinical spectrum of psoriatic patients in order to generate validated methods of assessment, particularly with regards to the prognostic ability of BIA and PhA. Further studies are needed to address the discrepancies in bioimpedance parameters within body compartments and between different devices and the deviation from health to disease. We hope that future studies will reveal insights into drug-specific alterations in body composition profiles in psoriatic disease, enabling clinicians to practice more stratified medicine and treat more effectively the metabolic components of patients’ disease that are so often neglected in clinical practice and associated with worse outcomes.

